# A pattern shift in SARS-CoV-2 Omicron variant transmission after the city lockdown--observational study based upon daily reported addresses of infected cases

**DOI:** 10.1101/2022.09.02.22279556

**Authors:** Lihong Yin, Guozhou Zhang

## Abstract

**Background:** Varied degrees of lockdown have been imposed for dozens of jurisdictions upon facing the SARS-CoV-2 epidemics during the past two years. Areal lockdown has been demonstrated effective to reduce the morbility and mortality of COVID-19. Even after the strict lockdown the peak of infection will appear around 9-25 days (median 18 days) thereafter. A wave of Omicron variant (BA.2 and BA.2.2) outbreak was seen from March to May 2022, in Shanghai, a megacity in China mainland. Aim To understand the sources of infection cases from outside or inside the isolated locations before and after the strict city lockdown.

**Methods:** The attributable addresses of SARS-CoV-2 infection were reported daily as well as the infected cases from March 18^th^, 2022 on through government website, which was publicly accessible. The address data and infected cases were collected until May 29th, 2022. The location (longitude and latitude) of these addresses were retrieved and the pattern of repeatedly reported addresses were analyzed. A tool of simple and meso-scale point-based (location-based) chronological graph was used to visualize and analyze the interactions of these locations.

**Results:** From March 18^th^ to May 29^th^ 2022, 173,350 items representing 35,743 unique addresses and 636,279 infected cases were released. The infection cases peaked 16 days after the city lockdown and were highly clustered in much crowded districts. The proportion of repeatedly reported locations of the previous day increased from around 20% before lockdown to greater than 40% in the plateau and remained at this level for up to one third (20/62) of the lockdown phase. This significantly increased proportion of intra-address infection indicated a pattern shift from inter-addresses to intra-address (D=0.2954, p<0.0001), which might perpetuate to the growth of infection cases. Based upon the day-to-day nearest neighbour transmission assumption the connections between some frequently repeated locations might be complex and heterogeneous.

**Interpretation:** During the strict areal isolation the intra-address infection may contribute significantly to infected cases of SARS-CoV-2 Omicron variant, the infection might have easily spilled over the boundary of family(with averaged family size of 2.3-3.1 people and family were required stay-at-home compulsively). This significant inter-addresses to intra-address pattern shift necessitated the understanding of intra-location transmission routes and corresponding interventions. Areal isolation and close off with homogeneous assumption inside and outside the isolated areas should be modified and the quantifing of the elevated risk for previously less exposed but much vulnerable sub-population was in pressing need.

**Research in context:** Evidence before this study It is commonly observable that the infection cases will continue to increase and peaked in several weeks after the first day of imposing areal lockdown. The search syntax of [(SARS-CoV-2 OR COVID-19) AND (transmission OR infection) AND (post-lockdown OR “during lockdown” OR “isolated area*”)] on PubMed hits 1372 records(on 2022-08-10). However, the viral transmission between the isolated locations or within them have not been studied thoroughly, maybe, due to the presumption that it would be stopped eventually by greatly reducing the inter-personal contact.The search syntax of [(SARS-CoV-2 OR COVID-19) AND (transmission OR infection) AND (post-lockdown OR “during lockdown” OR “isolated area*”) AND (strict)] on PubMed hits 57 records. The populations of different ages with heterogeneous risks of exposure to viruses have been revealed.

**Added values in this study:** The strict city lockdown of Shanghai in facing the Omicron outbreak provided a prototype to understand the sources of infection by using the daily reported address (isolated immediately) involving infected cases. A noticeable pattern shift was revealed in this study with significantly increased proportion of intra-location (small area) self-propagation after the city lockdown.

**Implication of all the available evidence:** Counter measures should be provided to reduce the risk of transmmision within the small isolated areas; and the elevated risk for the much vulnerable sub-populations within the intra-location (small isolated areas) should be quantified and to trade off between the reduced inter-location risk and increased intra-location risk of infection in policy making.

## Introduction

The lockdown had been hurriedly implemented to mitigate the spread of the severe acute respiratory syndrome coronavirus-2 (SARS-CoV-2) in dozens of jurisdictions since the city lockdown of Wuhan from the February 4, 2020 on[1-4], largely due to the lack of the vaccine and few of tested pharmaceuticals ever before were available to prevent and treat the infected cases effectively. The areal lockdown as indicated in measures of the Oxford COVID-19 Government Response Tracker (OxCGRT) would reduce the inter-personal contact, restrict the individuals movement and gathering and, in some extreme cases, would require to stay at home[5]. Varied degress of areal lockdown, therefore, had been demonstrated effective in slowing down the pace of transmission and/or reduction of mortality in facing single wave of COVID-19 attack in the compartment-based transmission dynamic modeling or in reality[4,6-12]. Under the constant threat of infection of evolving SARS-CoV-2 virus the easing of lockdown measures (re-opening) may be followed by another wave of infection or another period of lockdown in some jurisdictions[2,3,13] without adaptive post-lockdown disease control framework[14,15].

After the strict lockdown the peak of infected cases would appear 9-25 days (median 18 days) after the first day of imposing lockdown measures at the province level, and in this range the more heavily affected regions or the more reduciton in the mobility(with individual’s mobility reduction of ≥50%), the sooner to reach the peak[16]. During the lockdown period in Denmark the estimated secondary household infection rate was 17% (with basic and effective reproduction number of 2.1-4.7 and 2.5 for Alpha variant of SARS-CoV-2) and 82% of the secondary infection occured within 14 days[17,18]. Compared with the Delta variant of SARS-CoV-2 the Omicron variant indicated elevated basic reproduction number (with basic and effective reproduction number of 8.2 and 3.6)[19], shortened latent period (range 33-75 hours)[20]; and the vaccinated cases were expected to shed infectious virus for 6-9 days from onset of symptoms or diagnosis[21]. Therefore, it is reasonable to expect an incresed proportion of household secondary infection within shorter latent period for Omicron variant than its predecessors, as would be indicated by much steep trajectory curve of daily infected cases with elevated plateau during the city lockdown.

Due to the strict international border regulation, quarantine and close-circle-based management of relevent staffs China had almost completely prevented the reintroduction of SARS-CoV-2 variants of Alpha and Delta in past two years. Meanwhile, 86.1% of the Chinese people were full vaccinated (most majority with inactivated vaccine)[22,23] and had to adopt the routine physical distancing, face masking and hand hygiene (posted on 2020-11-22)[24]. Shanghai, a megacity in China, had adopted a precise strategy for isolation and close-off of the building (geographical address as proxy indicator) immediately with infected cases and quarantine of those close-contacts in the routine disease control. This building-based isolation ranged from close-off streetside shop to edifice, to resident quarter with several buildings or natural villages in the countryside. An epidemic of SARS-CoV-2 Omicron variant (BA.2 or BA.2.2) broke out in Shanghai since March 2022, and from March 29 on, the city were completely locked down in a two-step appraoch in two days[25]. The infected cases surged after the city lockdown and these phenomena were also seen in many previous jurisdictions imposing lockdown. Transmission dynamics and the effectiveness of lockdown were studied in some jurisdictions[1,3,5,6,9,15,18,26-29], where these infected cases come from remain largely unexplored. In this study, we collected the daily confirmed infected cases as well as their attributable address data from March 18 to May 29 2022, and analyzed the patterns of points in a day-to-day basis. The study aim is to reveal the pattern of address of infected cases before and after city lockdown and to understand the latent interactions between these points. An increased portion of intra-location infections were reported after the strict city lockdown.

## Data and methods

### Data

The isolated addresses in each district of Shanghai (China mainland) with SARS-CoV-2 infection cases were daily announced through the website of Shanghai Municipal Health Commission[24] since the March 18^th^, 2022. These data also with the infection cases released daily were publicly accessible. The confirm of the infected cases, the timeline of municipal public response and the arrangement of repeated massive screening tests for the nucleic acid of SARS-CoV-2 virus by using pharyngeal swab could refer to a descriptive study[25]. The longitude and latitude (in bd09 format) of these addressed were retrieved through web-based tool[30]. The addresses with missing longitude and latitude were double-checked on Amap website[31] through API of registered developer. Then the longitude and latitude (in bd09 or gcj02 format) were converted to the WGS84 format through user-defined function (reference code from [32]).

The SARS-CoV-2 infection including clinical diagnosed cases and asymptomatic infection (not meet the Protocol for Diagnosis and Management of COVID-19, 9th version in Chinese). All the positive SARS-CoV-2 cases were confirmed in officially accredited laboratory by using quantitative reverse-transcriptase-polymerase chain-reaction (qRT-PCR).

The map data of Shanghai were got from Aliyun website[33] in GeoJSON format was converted to shapefile through online tool[34] (WGS84 format). The Shanghai population of 7th census (on 2020-11-01) and area of district were retrieved from government portal website[35].

### Methods

The raw address data in each administrative district were stored into dataframe by attachment of date of release. Based upon the assumption of day-to-day nearest neighbour transmission, the date of address release in day 1 was used as reference in search of the nearest neighbour of the address of the following day (day 2, one day lag in this study). The nearest neighbour function (spatstat.geom::nnwhich) was used in point-wised search of the location from the day 2 to day 1[36], and the euclidean distance was calculated between this pair of points. All data manipulation and statistical analysis were conducted by using R(version 4.0.1)(details in Supplement materials B)[37]. As no individuals were identified this study could exempt from ethical review[38].

## Results

From the mid of March to the mid of May of 2022, Shanghai seen a surged wave of SARS-CoV-2 Omicron variant (BA.2 and BA.2.2) infection[39], meanwhile 627,775 cases were reported from March 6^th^, 2022 to May 29^th^, 2022[24]. The infection peaked 16 days after the city lockdown, and 22 days after the inflection point of cureve (Figure 1A). According to the data released of the national seventh census (2020-11-01) more than 24.87 million residents lived in this highly crowded metropolitan, which covered only 6,340.5 km^2^, with 3,392 people per km^2^. Still the population density in each of the district of city varied greatly, from around 500 people per km^2^ to greater than 32,000 people per km^2^. The infection was highly clustered in much crowded districts (Figure 1B, 1C). If the proportion of people per km^2^ infected were considered, the sixteen districts could be arbitrary divided into three subgroups, >5.0% (HP=9.3%), 2.5%-5.0% (PD, XH, HK, JA, YP, CN) and <2.5%(MH, BS, PT, JD, SJ, JS, QP, FX, CM) (Figure 1C). The infectious diseases tend to cluster because the people are clustered. The addresses of confirmed SARS-CoV-2 infection were released from March 18^th^, 2022 on, we collected 173,350 items of addresses from the government website until May 29^th^, 2022. After removing of the 50 addresses can not be located with longitude and latitude and other unrecognizable items the final analysis data set containing unique 35,743 addresses. The final location data set containing 172,843 addresses with longitude and latitude including 32,360 unique longitude and lattitude combined locations (up to 9.5% addresses were reported with different descriptions). Each address had reported about 17 cases on average (626379/35743) and had involved about five families in each address on average (estimated family size of 2.3-3.1 people in Shanghai). The reported locations of infection displayed pattern of human preferential design and/or of geographical features in this city, far from random spatial distribution (Figure 1D).

**Figure 1:**
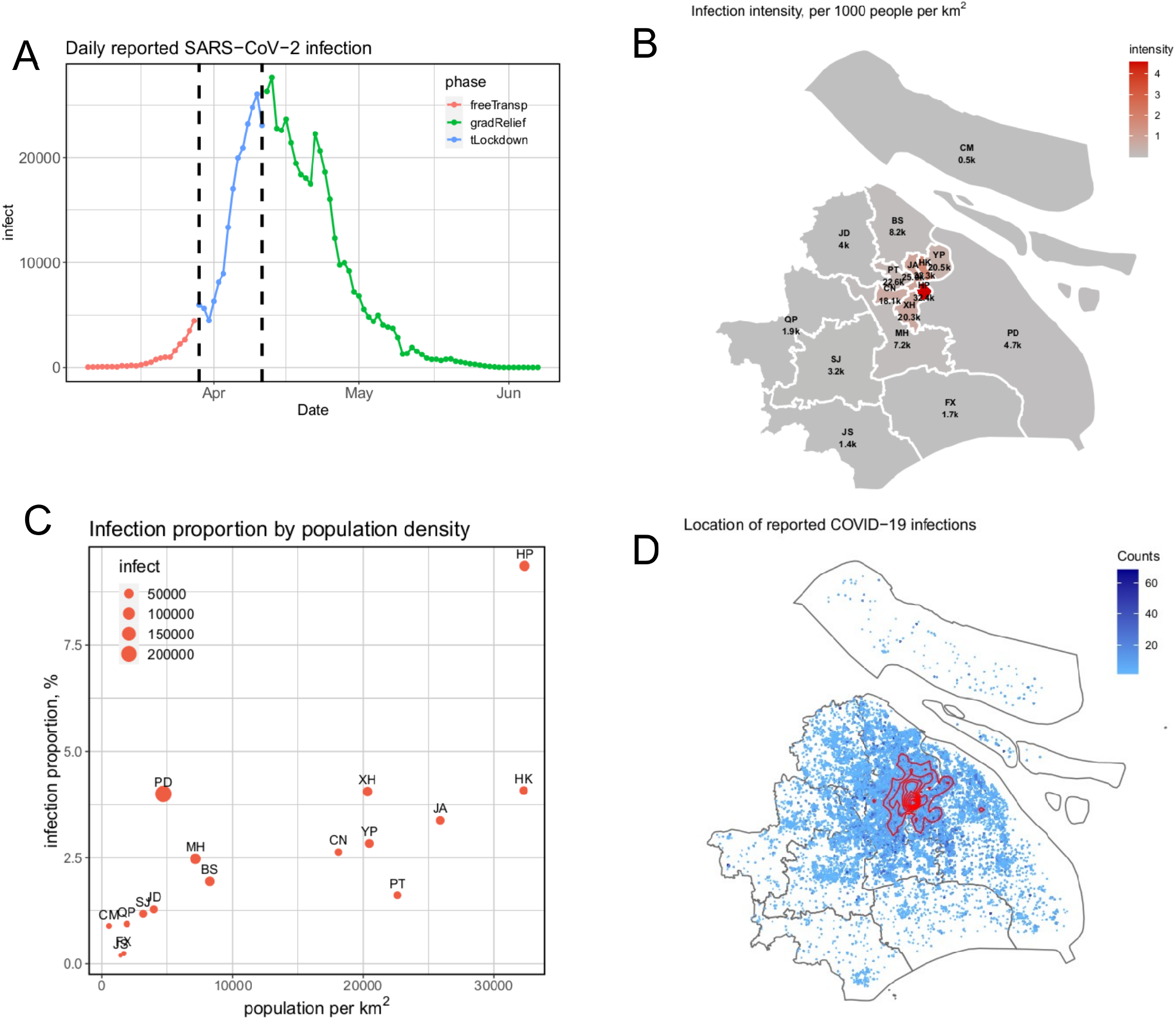
The epidemiological profile of SARS-CoV-2 Omicron variant (BA.2 and BA.2.2) in Shanghai (March 2022-May 2022 epidemics). A, daily reported cases of SARS-CoV-2 infection through public website; the metropolitan Shanghai gradually implemented the strict lockdown from Mar 29th, 2022 on; two phases division based upon this date were before and after city lockdown; alternatively, within the period of city lockdown the municipal government declared a three-tier classification and control of the addresses(isolated areas) based upon the time lapse from the date of detected infected cases or close contact, from April 11th, 2022, a period of gradient relief. B, the map of averaged infection intensity, namely, infected cases per 1000 people per km2 in each administrative district, the digits under the district name abbreviation indicate the population density per km2 (in kilo-person/km2). C, the proportion of infected cases in district population by the population density (people per km2); the dot size proprotion to the total infected cases in this study period; D, the reported locations with infected cases and the 2D density based contour line; the HP district was the hot area in this wave of infection.

We made an assumption that the single-reported infection location, by any reason, could be regarded as the terminal location of infection chain. Because the Omicron variant was highly contagious (with relative reproduction number of 2.71(95% CI 1.86-3.56) times of the Delta variant)[25,40,41], the probability of fail to transmit in these locations will be very low and negligible, especially when the virus-shedding infected cases have been there and stayed with colleagues or family members for longer than eight hours a day. This phenomenon may attribute to many reasons, including the very early detection and isolation of infected person (still in latent phase), implement of effective counter-measures including enough fresh air, physical distancing and timely eliminating of the residuals containing potential viruses, whatever the reason, the single-reported locations, to some extent, displayed the rapid and effective response to curb the viral spreading. On the other hand, the single-reported locations in some extreme cases would contain the single-deweller and/or the (the older) people who did not live at their own apartment, still the portion of each possible reason was unknown by now. We then checked the frequency of address reported before (upto 2022-03-28) and after (from 2022-03-29 onward) the lockdown of the city[25].

A pattern shift was seen following the city lockdown (from 2022-03-29 on), more locations appearred repeatedly, the proportion of single-reported address was higher before the city lockdown (two sided chi-square test, chi-square=1665, p<0.0001); however, the proportions of the repeatedly reported addresses increased following city lockdown (chi-square test for goodness-of-fit, chi-square=1274, p=0.0005) (Figure 2A, 2B). The repeated addresses were statistically increased after the city lockdown in two phases (Kolmogorov-Smirnov test, D=0.2954, p<0.0001)(Figure 2A) or three phase division (Kolmogorov-Smirnov test, D=0.1594, p<0.0001) (Figure 2B). The proportion of repeated locations of the previous day increased as the daily reported locations growth, increased from around 20% before lockdown to greater than 40% in the plateau and remained at this level for up to one third (20/62) of the lockdown phase (Figure 2C). Based upon the averaged infection cases per address in each day the increased proportion of repeated addresses would contribute increasingly (>40%) to the daily infected cases in the lockdown phase. The single-reported locations (n=13845) accounted for 38.7% of all addresses in final analysis data set, dramatically reduced from 68.1% of pre- to 41.7% of post-lockdown, also they displayed the feature of cluster (Figure 2D). Based upon the same infectious potential of virus, this obvious pattern shift of viral transmission from inter-addresses to the intra-address may explain a large part of why the infected cases yet surged even the whole city had been strictly locked down. After the specific locations had been locked the infection within these locations did not stopped accordingly. Further data (not available) to compare the infected cases of inside or outside of each reported address could also support this inference. Next, we compared the point pattern of a randomly sampled subset (n=1000) of these single-reported addresses to those of the Monte Carlo simulations of the completely spatial randomness[36,42]. The single-reported addresses indicated features of clustered events both in empty space function (spatstat.core::Fest) and in the nearest neighbour distance distribution function (spatstat.core::Gest) (Figure 2E, 2F). The Clark-Evans test of aggregation also indicated the significant clustering (R=0.2329, p=0.001).

**Figure 2:**
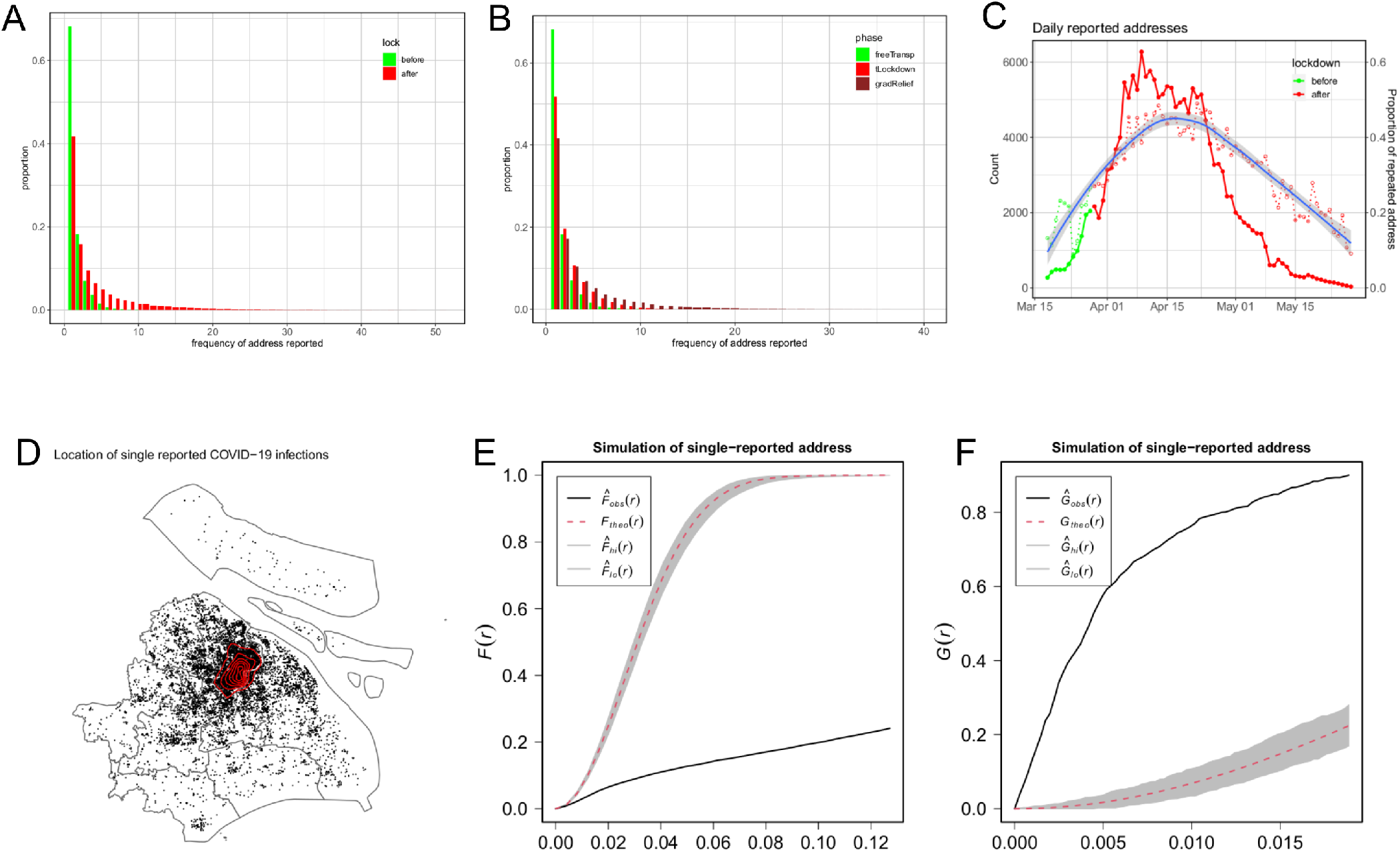
The proportion, growth and distribution of single reported locations. A, the proportion of single reported locations before and after city lockdown; B, the proportion of single reported locations before the city lockdown (free-Transp), totally lockdown (tLockdown) and gradient relief (gradRelief); C, the count of daily reported addresses (solid dot and left side y-axis) and the proportion of repeatedly reported addresses of the previous day (empty dot and right side y-axis); the blue line of fitted curve indicated the loess smooth and its 95% confidence interval of the proportion data. D, the actual distribution of single reported locations; E, the enveloped results of the comparison of the F function of a random sample of 1000 single reported locations with the 1000 times Monte Carlo simulation of complete spatial randomness; F, the enveloped results of the comparison of the G function of a random sample of 1000 single reported locations with the 1000 times Monte Carlo simulation of complete spatial randomness;

Before the city lockdown, the infected individuals with open social activity and free transportation were quite possible to display a long spatial distance transmission from some hubs, while the strict lockdown prohibited people to leave their home or their temporarily inhabiting site (e.g., staffs of essential social functions). Therefore, the spatial distance between sequential cases would be shorteded after lockdown. The reported locations of infection were highly clustered due to the high infection potential of virus through the easiest ways, to be convenient, in this study we made a second assumption that the individual might get infected in the shortest spatial distance from the location of previous cases including intra-location self-propagation (Figure 3A-3C). Though the incubation period span 1-14 days in natural course (median incubation of 3 days for Omicron variant)[20,43], part of infected cases in the following day could be viewed as the secondary infection of the same location especially when this location had been closed off and isolated from others for longer than the usual viral shedding period (6-9 days)[21]. Therefore, even lacked the individual’s trajectory data and could not be associated with the generations in the infection chain the repeatedly reported locations indicated high probability of the intra-location infection (endemic). Before the city lockdown, a small portion of locations displayed extended nearest neighbour distance(Figure 3D, 3E). This was quite possible attributed to the distance of commute. The proportion of intra-location secondary infection (location repeatedly reported ≥ 10 days) after the city lockdown (11.8%) was significantly higher than that of the before (0.097%), with the odds ratio of 138.7 (95% CI 59.3-432.3)(p<0.0001). Hitherto, the location data has displayed that the more locations had been reported repeatedly after the city lockdown, and a greater portion were repeated longer than the usual viral shedding period (6-9 days). We could learn that the location close off and isolation did not stop the infection within the location accordingly, spatially or in name.

**Figure 3:**
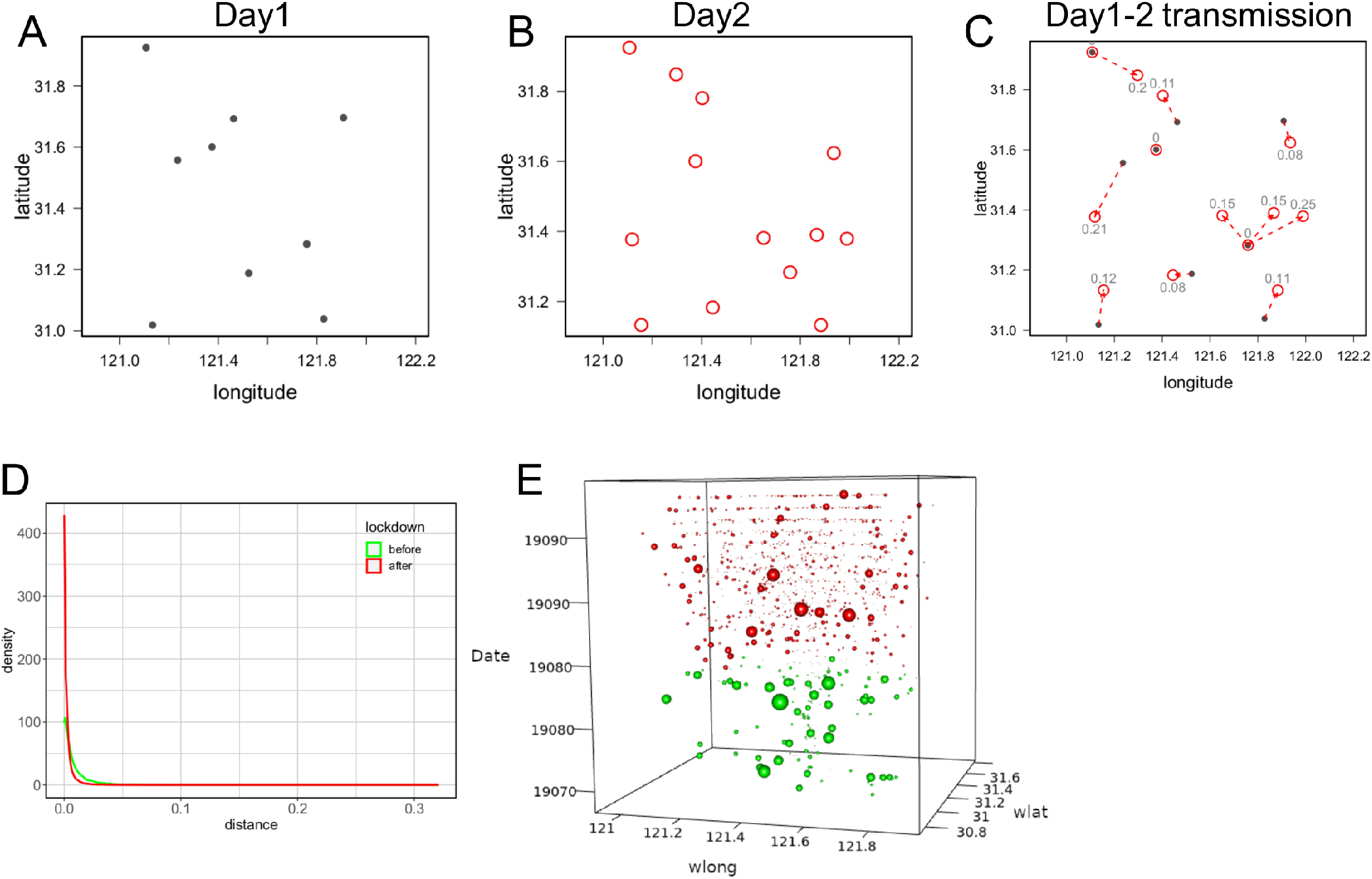
The point-wise viral spreading based upon the day-to-day nearest neighbour assumption. A-C, represent the toy data of point-wise day 1, day 2 and day-to-day viral transmission assumption; the point-wise nearest neighbour distance from the day 1 to day 2 was estimated by set the day 1 locations of reference as a whole and search the nearest neighbour of the locations of the day 2 point-by-point; D, the distribution of the nearest neighbour distance, the distance from the repeated address in second day to that of previous day was zero; E, visualize of the spatial distribution of nearest neighbour distance with sphere radius proportion to the estimated distance in the periods of before lockdown (green sphere for freeTransp) and city total lockdown (red sphere for tLockdown);

An increased portion of locations (>40%) repeatedly had reported after the city lockdown (Figure 2C, 4A), and these surged number of repeated locations would negate the validity of city lockdown if the intra-location transmission were not stopped (Figure 4B). To explore the transmission patterns of intra- and inter-locations, we incorporate the distance and temporal data into a point-based chronological network (Figure 5A-5G)[44]. It was possible to examine the pattern of transmission of each repeatedly appeared location based upon the regular time lag and a connection rule (one-day lag and the nearest neighbour distance transmission assumption in this study). As a result, there were 22,912 locations appeared as the source of previous day and 32,316 locations as the target of following day, they formed 172,584 location pairs in the final analysis dataset. By using this point-based spatio-temporal graph model we examined the some frequently appeared locations. Based upon the day-to-day nearest neighbour transmission assumption the infection chain turned out to be complex connected, and the intra-location infection might have perpetuated the chain (Figure 6A,6B). It was also noticeable that as the infection cases increased from before to after the city lockdown the inter-locations transmission become more complex. This point-based spatio-temporal graph could be applied in further scenarios(Figure 6C, 6D). Some pivotal locations and time-point were also detectable (Figure 7, Figure 8). These pivotal points might provide important clue to further control the disease spread even in the period of city lockdown(further verifying study required).

**Figure 4:**
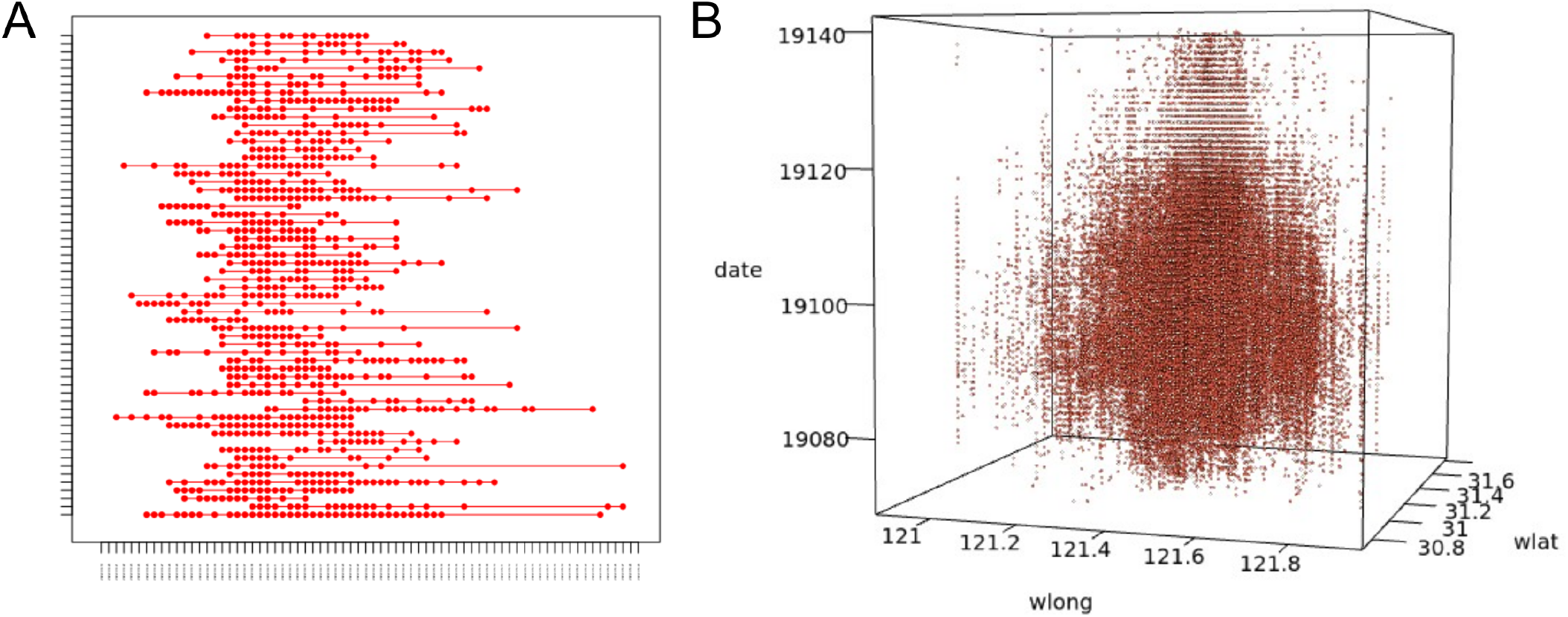
Example of pattern of repeatedly reported locations. A, Example of the repeated locations along the date; B, Examples of the repeated locations along the date with longitude and latitude;

**Figure 5:**
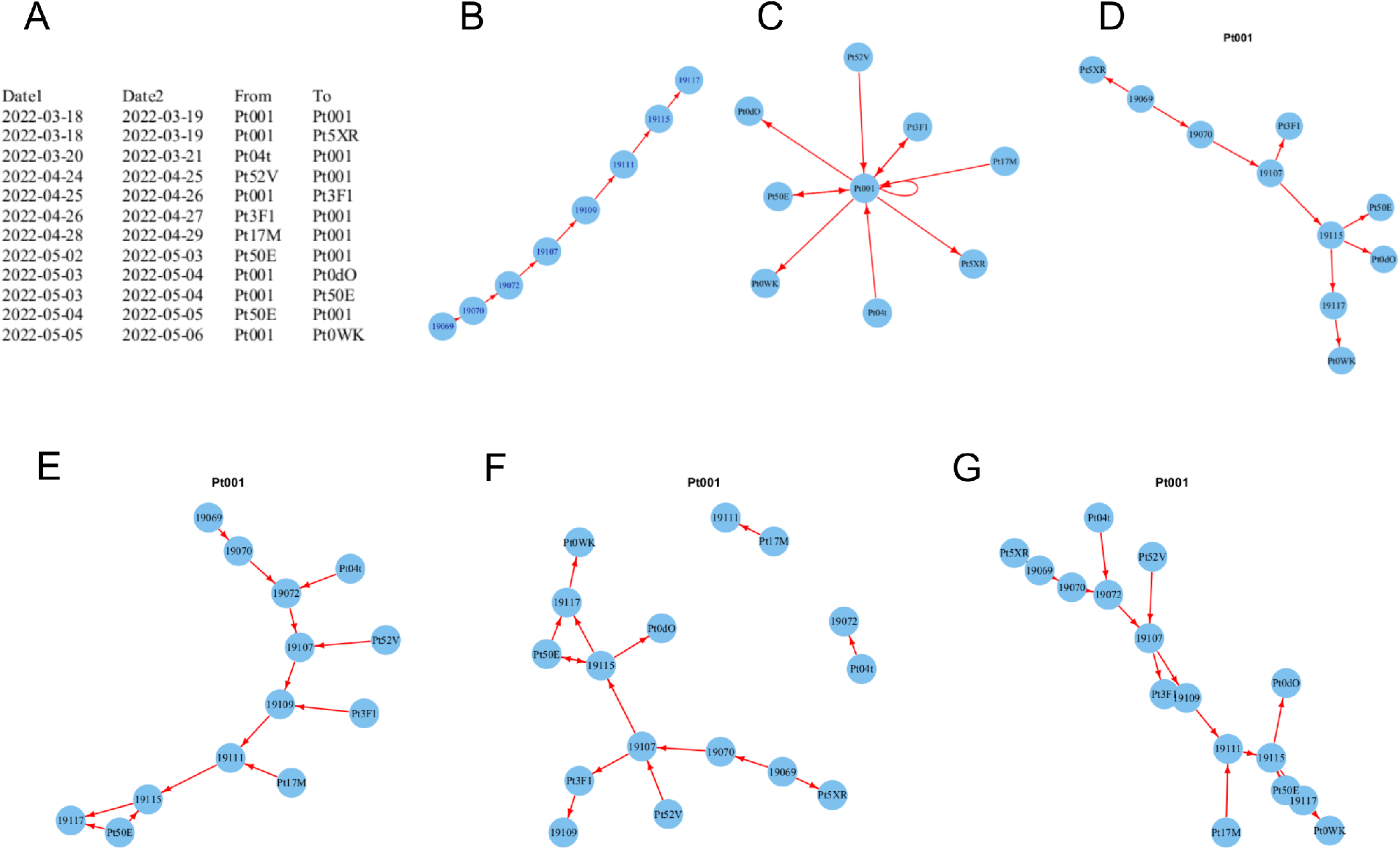
To generate directed acyclic graph by combining spatio-temporal data. A, a toy dataset; B, chronological sequence of specific point and the date was converted into integer dated from the origin of 1970-01-01; C, the directed acyclic graph of specific point; D, graph combining specific point as source of transmission and its chronological sequence (source chain); E, graph combining specific point as target of transmission and its chronological sequence (target chain); F, graph combining specific point both as source and target of transmission and its chronological sequence (source chain plus target points); G, graph combining the chronological sequence and both as source and target of transmission (full chain);

**Figure 6:**
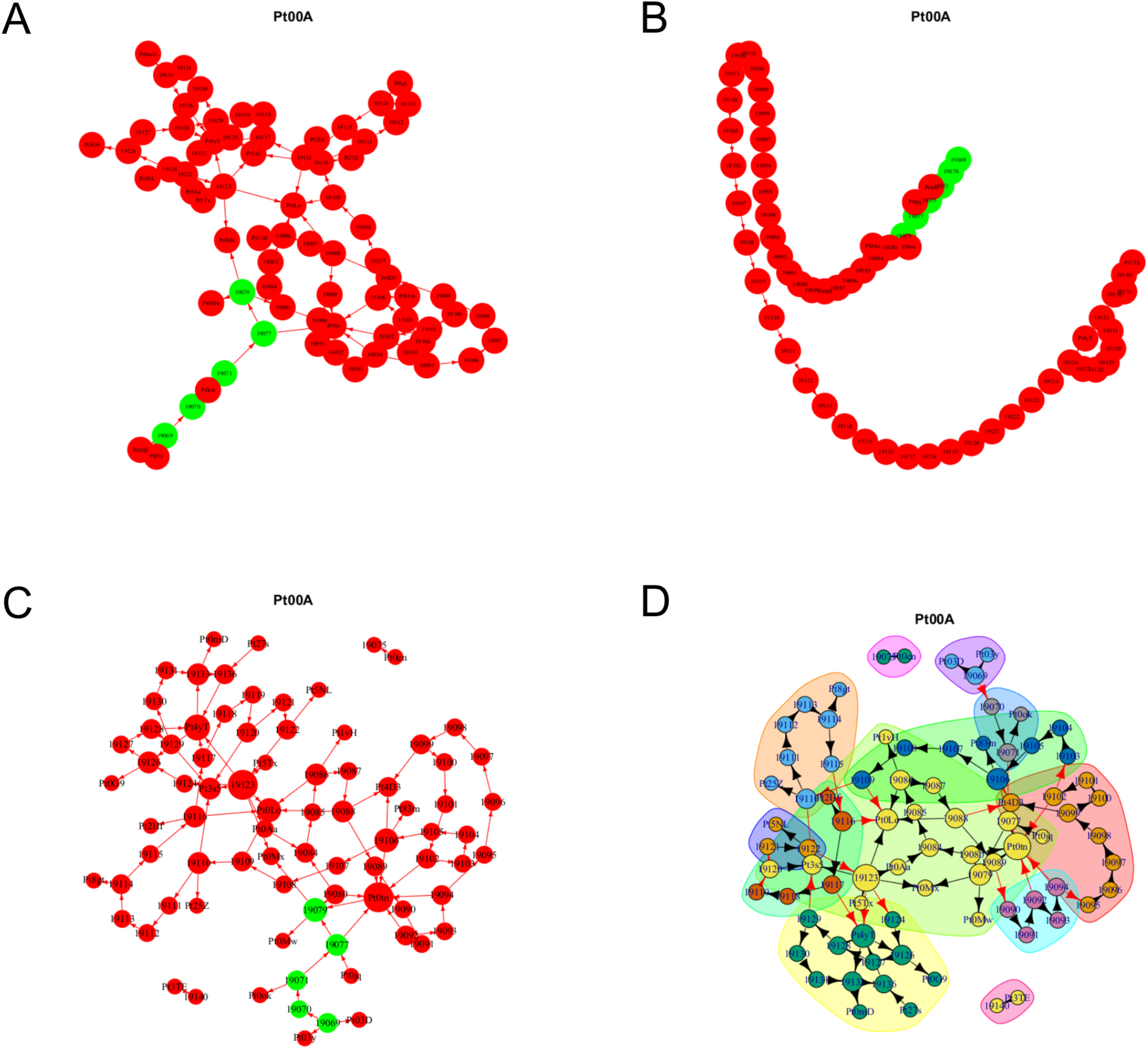
Point-based chronological graph of one frequently repeated location. A, source chain graph of the point Pt00A; B, target chain graph of the point Pt00A; C, source chain plus target points of the point Pt00A modified by rescale the vertex size by degree of connection; D, full chain of the point Pt00A clustered bycommunity (connection density) via random walk;

**Figure 7:**
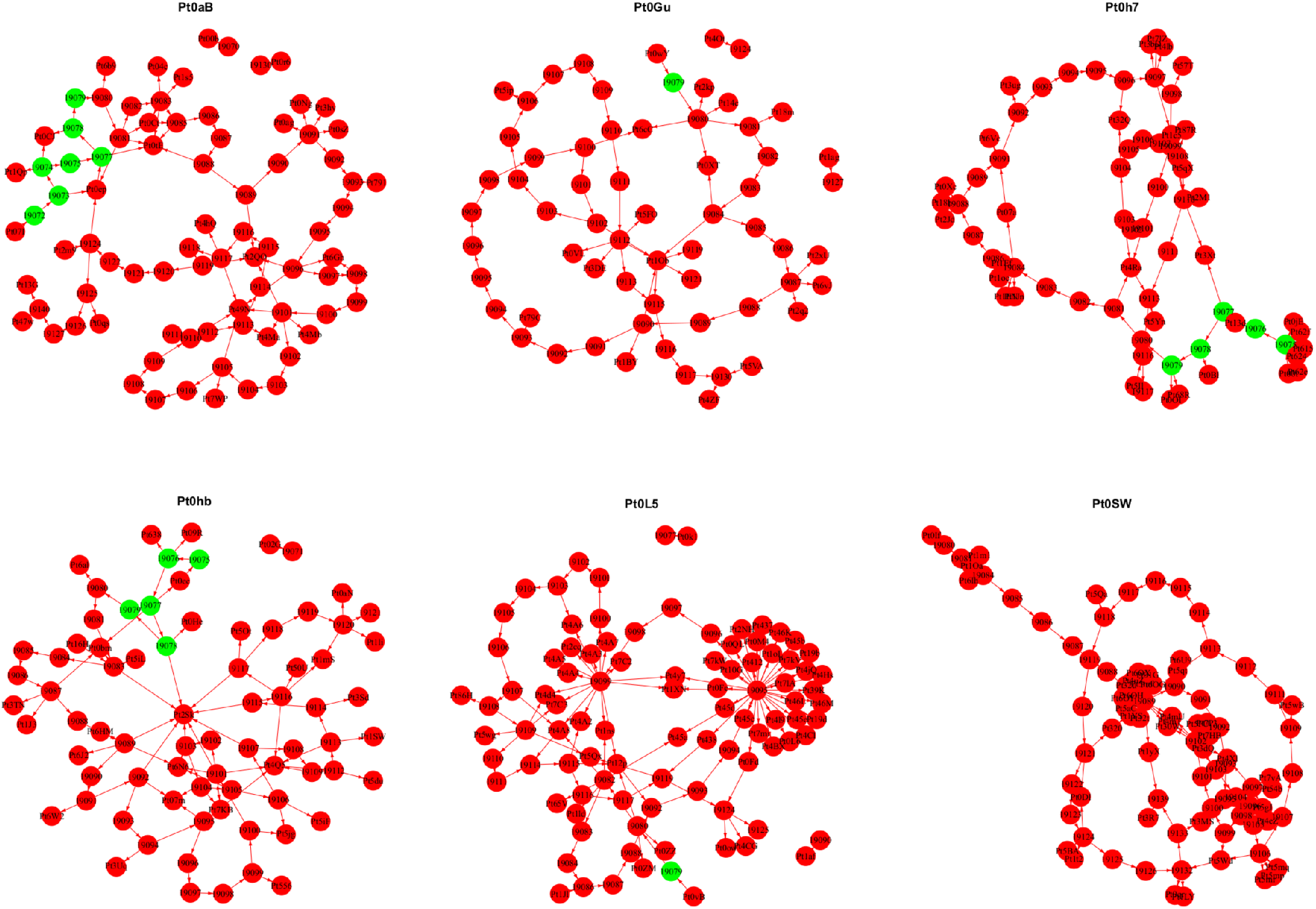
Some commonly appearred locations as source based upon the day-to-day nearest neighbour transmission assumption (green and red nodes represented date before and after the city lockdown, respectively).

**Figure 8:**
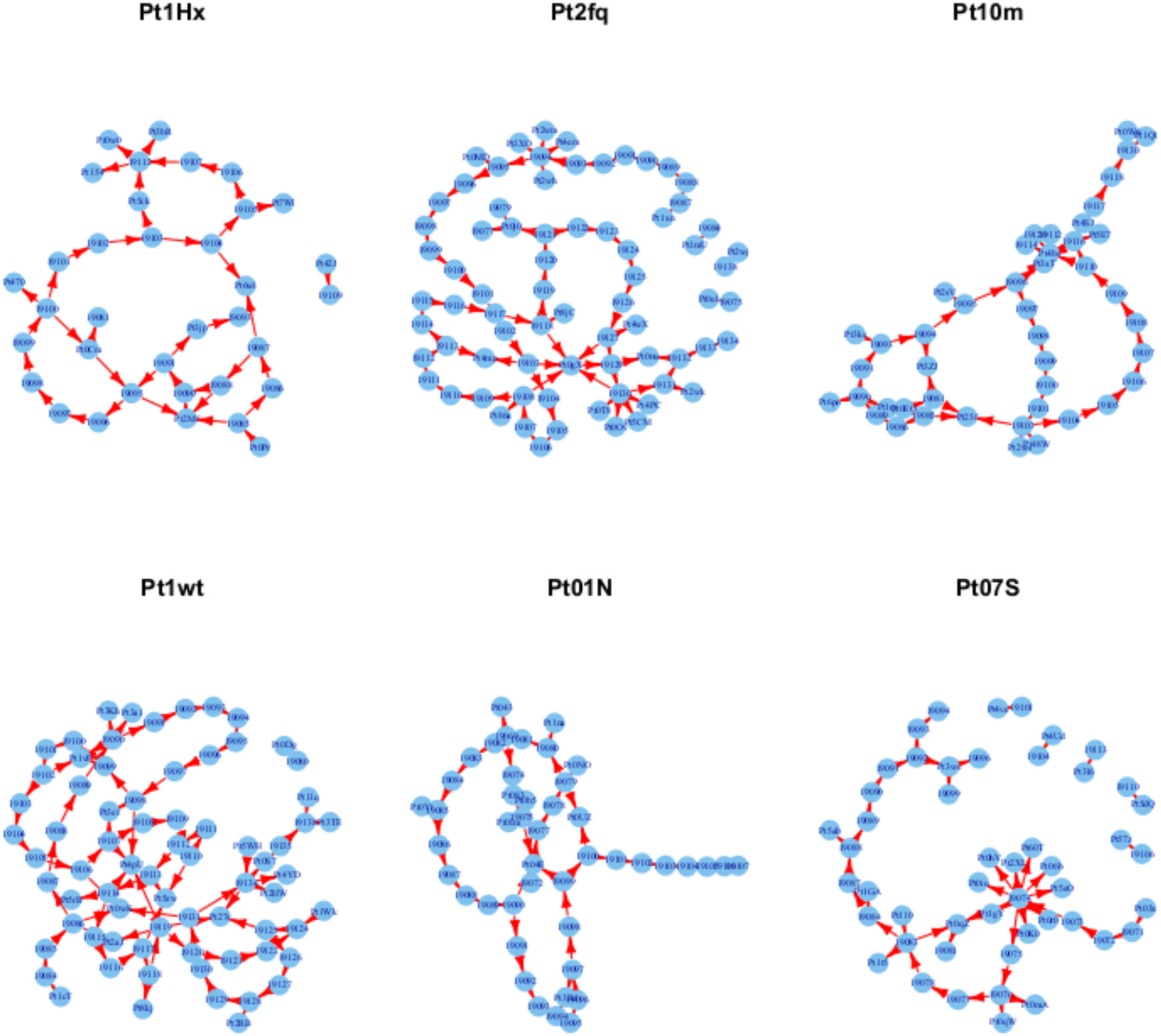
Some commonly appearred the locations as target based upon the day-to-day nearest neighbour transmission assumption.

Next, we conducted pair-wise graph morphism comparison between those locations with the same times of appearance as presumed source of infection (Supplement Figure s01-s04). We arbitrarily selected 756 locations with the same times of appearance as presumed source of infection, 52,102 pair-wised graph morphism comparisons failed to find the isomorpic graphs (data not presented). The cluster analysis of the locations in the sequence of date of appearance did not provide common patterns (Supplement Figure s05). The results suggested that the viral transmission inter- and intra-locations might be complicated as the underlying routes had not been fully understood yet.

## Discussion

The SARS-CoV-2 Omicron variant had demonstrated its high contagious once again from the mid of March to the mid of Jun 2022 in a megacity Shanghai, China. We had seen the growth of the infected cases even after the whole city had implemented the strictest lockdown policy (from 2022-03-29 on). The viral infection were clustered due to people are gathered, the most crowded district suffered from the highest proportion of infection correlated with the its population density. As the daily infected case increased the proportion of the repeatedly reported addresses increased accordingly and brought a significant pattern shift of the proportion profile from before to after the city lockdown. The intra-location infection might account for greater than 40% of all infected cases during the city lockdown phase. Considering each unique address had reported around 17 infected case on average, the transmission had spread over the boundary of family in the isolated addresses. During the strict city lockdown the residents in Shanghai were required to stay-at-home completely, therefore, the routes of transmission within isolated buildings with crowded residents should be explored. The lockdown would alter the pattern and behavoir of contact[45], however, the risk of susceptibility might be heterogeneous though not the same as before[46,47]. If the much vulnerable sub-population (e.g., the older >65 years of age) would face elevated risk of exposure the interfering mesaures within isolated address should be necessary[46]. Alternatively, a strategy to precisely pin down the boundary of transmission should be developed and implemented instead of arbitrary scope areal based isolation or broad scope lockdown though one might argue about the net gain of reduced infection cases during lockdown.

The decreased the proportion of the single-reported addresses (viewed as the terminal of infection chain) as the infected cases rocketted up indicated that the capacity of the effective counter-measures to control the spread of virus might have been saturated and been overwhelmed. If it’s true then the more practical policies and measures should be considered in high priority in response to such a large scale of the public emergency. On the other hand, a noticeable portion of addresses (>60%) newly appeared after the city lockdown though the city lockdown might have greatly reduced human movement and flatten or shorten the curve of of viral free infection[1,11,12,48,49]. It means that even under the strict lockdown the essential functions to support an ordered megacity is not a trivial, the electricity, water and gas supply, the hospital and police station, the logistics system, environmental workers, government, volunteers to fill in the posts in vacancy and many others. Therefore, we are facing a Corona dilemma(trolly problem) whether to exchange the more infections among low risk sub-population outside the isolated areas with the less infections among the more vulnerable older sub-population in the isolated areas. Areal isolation and close off like what had done in Shanghai in response to this wave of outbreak might be quite possible to result in the more older people facing elevated risk of exposure and infection[46], especially in a “grey hair” city. Further studies to quantify the modified risk of exposure and its consequences to different age sub-populations would be helpful for policy making.

We had assumed residents in these daily appeared new addresses had been infected from their spatially nearest neighbour(s). This was quite possible especially after the strict city lockdown during this period even the delivery of the materials to support basic living in ordinary residents had been minimized. The residents ordered their basic living materials once a week and the goods was delivered to the door by volunteers within the address under proper protection. It suggested that a large portion of residents and their supportive logistics system were localized within or near an isolated small area (address as proxy indicator). Among our location retrieval from the original address, 75%(32153/42948) indicated bias of 100-300 meters, the other 25%(10793/42948) indicated bias of 1-10 kilometers[50]. These centroid locations represented identifiable areas numbered down the road or street and villages in the countryside. In quantitative spatial inference locations with different precision should be treated differently, we largely exempted this problem because the location only be used in estimating the distance from its nearest neighbours and this distance would be the same before and after the city lockdown. To generate the point-based chronological graph[44] (based upon the day-to-day transmission assumption in this study) the nearest neighbours were used to demonstrate the concept. Other rules could be used to generate the connection between these spatial points, especially when the vaccination, physical distancing, face masking, hand washing, early case detection (nucleic acid test) and prolonged viral shedding might had altered the dynamics of transmission of SARS-CoV-2[49,51-54]. We need more tools to understand the underlying routes of transmission. We provided a method to reveal the latent connection rather than to make inference from these location data in this study though further verifying studies would be required.

To be convinent we used the daily reported address directly to generate one day lag disease transmission graph, this means the infection generation (doubling time) was about 24 hours. This had not been demonstrated but was quite close to the actual data of Omicron variant transmission, 1.28 days in a community in Hong Kong[55]; or the latent period ranged from 33 to75 hours[20]. Aimed to keep pace with the transmission of Omicron variant an ambitious strategy had been proposed and implemented to run the relevent human resources at its full speed. Therefore, during the response to this wave of outbreak the cases newly infected, recovered and deceased or the newly involved addresses were reported daily and, the Center for Disease Control and Prevention (CDC) were demanded to finished the retropective records of the exposure history of infected cases within the day of newly cases were confirmed. We generate the point-based chronological graph by using lags of one to three days, the united graph would provide more informative clue (Supplement Figure s06).

The trajectory data of individuals had been used to track potential exposure and the infection[5,9,16,29,48,56], however, these data were not accessible for the CDC work force and reliable models to identify the persons of potential exposure might be developing. The point-based chronological graph could be viewed as a meso-scale method to visualize and analyze the connectivity (base upon some rules) and interactions between spatial locations. It was easy to generate and simple to manipulate by using the graph tools, but it could not be applied to predict without conditional probabilities such as those in Bayesian network. Still, we are under the threat of constant evolving SARS-CoV-2 viruses and the lockdown of different scope jurisdictions are ongoing especially in those areas with a large proportion of much vulnerable people.

## Conclusion

We had seen a wave of outbreak of SARS-CoV-2 Omicron variant from March to the end of May 2022 in Shanghai, a megacity of China mainland. By using the publicly accessible data released daily during this period we revealed a pattern shift of the addresses of infected cases. After the city lockdown an elevated proportion of address were reported repeatedly, which indicated an increased proportion of the intra-location viral transmission. A simple and meso-scale tool that can adopt different rules to chronologically visualize and analyze the connectivity and interactions may be useful for infection control.

## Data Availability

https://wsjkw.sh.gov.cn/xwfb/index.html

## Authorship contribution statement

Lihong Yin: design the study, collect and analyze the data, drafting; Guozhou Zhang: design the study, collect and analyze the data, drafting;

## Date sharing

All raw data are publicly accessible through websites.

## Acknowledgements

We are gratful to all the people facing this wave of Omicron variants outbreak in Shanghai, including general residents who have to stay-at-home, working force who have to shoulder extra responsibilities and risk of infection to maintain the essential functions of the city, and who have provided support of any kind from inside or outside.

## Supplementary material

A supplement material is available including the figures, detailed library information used in data analysis and source code of user-defined functions in this study.

## References

1. Sun GQ, Wang SF, Li MT, et al. Transmission dynamics of COVID-19 in Wuhan, China: effects of lockdown and medical resources. Nonlinear Dyn. 2020;101(3):1981–1993. doi:10.1007/s11071-020-05770-9

2. Anderson SC, Mulberry N, Edwards AM, et al. How much leeway is there to relax COVID-19 control measures? Epidemics. 2021;35:100453. doi:10.1016/j.epidem.2021.100453

3. Nadim SS, Chattopadhyay J. Occurrence of backward bifurcation and prediction of disease transmission with imperfect lockdown: A case study on COVID-19. Chaos Solitons Fractals. 2020;140:110163. doi:10.1016/j.chaos.2020.110163

4. Hsiang S, Allen D, Annan-Phan S, et al. The effect of large-scale anti-contagion policies on the COVID-19 pandemic. Nature. 2020;584(7820):262–267. doi:10.1038/s41586-020-2404-8

5. Santamaria C, Sermi F, Spyratos S, et al. Measuring the impact of COVID-19 confinement measures on human mobility using mobile positioning data. A European regional analysis. Saf Sci. 2020;132:104925. doi:10.1016/j.ssci.2020.104925

6. Bugalia S, Bajiya VP, Tripathi JP, Li MT, Sun GQ. Mathematical modeling of COVID-19 transmission: the roles of intervention strategies and lockdown. Math Biosci Eng. 2020;17(5):5961–5986. doi:10.3934/mbe.2020318

7. Flaxman S, Mishra S, Gandy A, et al. Estimating the effects of non-pharmaceutical interventions on COVID-19 in Europe. Nature. 2020;584(7820):257–261. doi:10.1038/s41586-020-2405-7

8. Verma BK, Verma M, Verma VK, et al. Global lockdown: An effective safeguard in responding to the threat of COVID-19. J Eval Clin Pract. 2020;26(6):1592–1598. doi:10.1111/jep.13483

9. Vinceti M, Balboni E, Rothman KJ, et al. Substantial impact of mobility restrictions on reducing COVID-19 incidence in Italy in 2020. J Travel Med. Published online July 24, 2022:taac081. doi:10.1093/jtm/taac081

10. Pleninger R, Streicher S, Sturm JE. Do COVID-19 containment measures work? Evidence from Switzerland. Swiss J Econ Stat. 2022;158(1):5. doi:10.1186/s41937-022-00083-7

11. Stokes J, Turner AJ, Anselmi L, Morciano M, Hone T. The relative effects of non-pharmaceutical interventions on wave one Covid-19 mortality: natural experiment in 130 countries. BMC Public Health. 2022;22(1):1113. doi:10.1186/s12889-022-13546-6

12. Venkatesh U, Gandhi P A, Ara T, Rahman MM, Kishore J. Lockdowns, Community Mobility Patterns, and COVID-19: A Retrospective Analysis of Data from 16 Countries. Healthc Inform Res. 2022;28(2):160–169. doi:10.4258/hir.2022.28.2.160

13. Day M. Covid-19: Italy reimposes widespread lockdown as transmission rate rises again. BMJ. 2021;372:n726. doi:10.1136/bmj.n726

14. Ikoona EN, Kitara DL. A proposed framework to limit post-lockdown community transmission of COVID-19 in Africa. Pan Afr Med J. 2021;38:303. doi:10.11604/pamj.2021.38.303.24008

15. Frost I, Craig J, Osena G, et al. Modelling COVID-19 transmission in Africa: countrywise projections of total and severe infections under different lockdown scenarios. BMJ Open. 2021;11(3):e044149. doi:10.1136/bmjopen-2020-044149

16. Vinceti M, Filippini T, Rothman KJ, et al. Lockdown timing and efficacy in controlling COVID-19 using mobile phone tracking. EClinicalMedicine. 2020;25:100457. doi:10.1016/j.eclinm.2020.100457

17. Boldog P, Tekeli T, Vizi Z, Dénes A, Bartha FA, Röst G. Risk Assessment of Novel Coronavirus COVID-19 Outbreaks Outside China. J Clin Med. 2020;9(2). doi:10.3390/jcm9020571

18. Lyngse FP, Kirkeby C, Halasa T, et al. Nationwide study on SARS-CoV-2 transmission within households from lockdown to reopening, Denmark, 27 February 2020 to 1 August 2020. Euro Surveill. 2022;27(6). doi:10.2807/1560-7917.ES.2022.27.6.2001800

19. Liu Y, Rocklöv J. The effective reproductive number of the Omicron variant of SARS-CoV-2 is several times relative to Delta. J Travel Med. 2022;29(3). doi:10.1093/jtm/taac037

20. Jansen L, Tegomoh B, Lange K, et al. Investigation of a SARS-CoV-2 B.1.1.529 (Omicron) Variant Cluster - Nebraska, November-December 2021. MMWR Morb Mortal Wkly Rep. 2021;70(5152):1782–1784. doi:10.15585/mmwr.mm705152e3

21. Takahashi K, Ishikane M, Ujiie M, et al. Duration of Infectious Virus Shedding by SARS-CoV-2 Omicron Variant-Infected Vaccinees. Emerg Infect Dis. 2022;28(5):998–1001. doi:10.3201/eid2805.220197

22. Li M, Liu Q, Wu D, et al. Association of COVID-19 Vaccination and Clinical Severity of Patients Infected with Delta or Omicron Variants - China, May 21, 2021-February 28, 2022. China CDC Wkly. 2022;4(14):293–297. doi:10.46234/ccdcw2022.074

23. WHO. WHO Coronavirus(COVID-19) Dashboard. WHO Coronavirus(COVID-19) Dashboard. Accessed August 5, 2022. https://covid19.who.int/

24. Shanghai Municipal Health Commission. News Release. Accessed March 1, 2022. https://wsjkw.sh.gov.cn/

25. Chen Z, Deng X, Fang L, et al. Epidemiological characteristics and transmission dynamics of the outbreak caused by the SARS-CoV-2 Omicron variant in Shanghai, China: a descriptive study. medRxiv. Published online June 18, 2022:2022.06.11.22276273. doi:10.1101/2022.06.11.22276273

26. Askar SS, Ghosh D, Santra PK, Elsadany AA, Mahapatra GS. A fractional order SITR mathematical model for forecasting of transmission of COVID-19 of India with lockdown effect. Results Phys. 2021;24:104067. doi:10.1016/j.rinp.2021.104067

27. Saeed U, Sherdil K, Ashraf U, et al. Identification of potential lockdown areas during COVID-19 transmission in Punjab, Pakistan. Public Health. 2021;190:42–51. doi:10.1016/j.puhe.2020.10.026

28. Yang HM, Lombardi Junior LP, Castro FFM, Yang AC. Mathematical modeling of the transmission of SARS-CoV-2-Evaluating the impact of isolation in São Paulo State (Brazil) and lockdown in Spain associated with protective measures on the epidemic of CoViD-19. PLoS One. 2021;16(6):e0252271. doi:10.1371/journal.pone.0252271

29. Beria P, Lunkar V. Presence and mobility of the population during the first wave of Covid-19 outbreak and lockdown in Italy. Sustain Cities Soc. 2021;65:102616. doi:10.1016/j.scs.2020.102616

30. Map Location. Address to Location. Accessed May 30, 2022. https://maplocation.sjfkai.com

31. Amap Web API. Amap Web API. Accessed June 20, 2022. https://lbs.amap.com

32. Yankai Wang. CSDN Blog--Basic dev-tools and configuration. Python map coordinates inter-convertion (bd09, gcj02, wgs84). Published April 14, 2021. Accessed June 20, 2022. https://blog.csdn.net/weixin_38468077/article/details/115689448

33. Aliyun Data Visualization Plotform. Datav.GeoAtlas. Accessed June 20, 2022. http://datav.aliyun.com/portal/school/atlas/

34. Mapshaper. Mapshaper. Accessed June 5, 2022. https://mapshaper.org

35. Shanghai Municipal Statistical Bureau. Shanghai Statistical Yearbook 2021. Accessed June 20, 2022. https://tjj.sh.gov.cn

36. Baddeley A, Turner R. spatstat: An R Package for Analyzing Spatial Point Patterns. Journal of Statistical Software. 2005;12(6):1–42. doi:10.18637/jss.v012.i06

37. R Core Team. R: A Language and Environment for Statistical Computing. Published online 2021. https://www.R-project.org

38. Ienca M, Vayena E. On the responsible use of digital data to tackle the COVID-19 pandemic. Nat Med. 2020;26(4):463–464. doi:10.1038/s41591-020-0832-5

39. Global Times. Shanghai’s latest outbreak caused by Omicron’s BA.2 and BA.2.2 mutations, no new variants detected.https://www.globaltimes.cn/page/202205/1265329.shtml. Published May 11, 2022.

40. Ito K, Piantham C, Nishiura H. Relative instantaneous reproduction number of Omicron SARS-CoV-2 variant with respect to the Delta variant in Denmark. J Med Virol. 2022;94(5):2265–2268. doi:10.1002/jmv.27560

41. Du Z, Hong H, Wang S, et al. Reproduction Number of the Omicron Variant Triples That of the Delta Variant. Viruses. 2022;14(4). doi:10.3390/v14040821

42. Baddeley A, Rubak E, Turner R. Spatial Point Patterns: Methodology and Applications with R. 1st ed. Chapman and Hall/CRC Press; 2015.

43. Boucau J, Marino C, Regan J, et al. Duration of viable virus shedding in SARS-CoV-2 omicron variant infection. medRxiv. Published online March 2, 2022:2022.03.01.22271582. doi:10.1101/2022.03.01.22271582

44. Ferreira LN, Vega-Oliveros DA, Cotacallapa M, et al. Spatiotemporal data analysis with chronological networks. Nat Commun. 2020;11(1):4036. doi:10.1038/s41467-020-17634-2

45. Bosetti P, Huynh BT, Abdou AY, et al. Lockdown impact on age-specific contact patterns and behaviours, France, April 2020. Euro Surveill. 2021;26(48). doi:10.2807/1560-7917.ES.2021.26.48.2001636

46. Lau MSY, Liu C, Siegler AJ, et al. Post-lockdown changes of age-specific susceptibility and its correlation with adherence to social distancing measures. Sci Rep. 2022;12(1):4637. doi:10.1038/s41598-022-08566-6

47. Arinaminpathy N, Das J, McCormick TH, Mukhopadhyay P, Sircar N. Quantifying heterogeneity in SARS-CoV-2 transmission during the lockdown in India. Epidemics. 2021;36:100477. doi:10.1016/j.epidem.2021.100477

48. Jia JS, Lu X, Yuan Y, Xu G, Jia J, Christakis NA. Population flow drives spatio-temporal distribution of COVID-19 in China. Nature. 2020;582(7812):389–394. doi:10.1038/s41586-020-2284-y

49. Zamir M, Shah Z, Nadeem F, Memood A, Alrabaiah H, Kumam P. Non Pharmaceutical Interventions for Optimal Control of COVID-19. Comput Methods Programs Biomed. 2020;196:105642. doi:10.1016/j.cmpb.2020.105642

50. Baidu. Baidu Map Open Service Platform. Baidu Map Web API. Published 2022. Accessed May 20, 2022. https://lbsyun.baidu.com/index.php?title=webapi/guide/webservice-geocoding

51. Anderson SC, Edwards AM, Yerlanov M, et al. Quantifying the impact of COVID-19 control measures using a Bayesian model of physical distancing. PLoS Comput Biol. 2020;16(12):e1008274. doi:10.1371/journal.pcbi.1008274

52. Tepekule B, Hauser A, Kachalov VN, et al. Assessing the potential impact of transmission during prolonged viral shedding on the effect of lockdown relaxation on COVID-19. PLoS Comput Biol. 2021;17(1):e1008609. doi:10.1371/journal.pcbi.1008609

53. Ma WJ, Wang XS, Tian H, et al. [Characteristics of SARS-CoV-2 Omicron infection in children imported from Hong Kong]. Zhonghua Er Ke Za Zhi. 2022;60(6):539–544. doi:10.3760/cma.j.cn112140-20220423-00367

54. Andrews N, Stowe J, Kirsebom F, et al. Covid-19 Vaccine Effectiveness against the Omicron (B.1.1.529) Variant. N Engl J Med. 2022;386(16):1532–1546. doi:10.1056/NEJMoa2119451

55. Cheng VCC, Ip JD, Chu AWH, et al. Rapid spread of SARS-CoV-2 Omicron subvariant BA.2 in a single-source community outbreak. Clin Infect Dis. Published online March 10, 2022. doi:10.1093/cid/ciac203

56. Zhang H, Yin L, Mao L, et al. Combinational Recommendation of Vaccinations, Mask-Wearing, and Home-Quarantine to Control Influenza in Megacities: An Agent-Based Modeling Study With Large-Scale Trajectory Data. Front Public Health. 2022;10:883624. doi:10.3389/fpubh.2022.883624

