## Supplemental Materials for "A pattern shift in SARS-CoV-2 Omicron variant transmission after the city lockdown--observational study based upon daily reported addresses of infected cases" (from another version of this preprint: version 1, 2022-09-04)

**Supplement A: supplement figures**

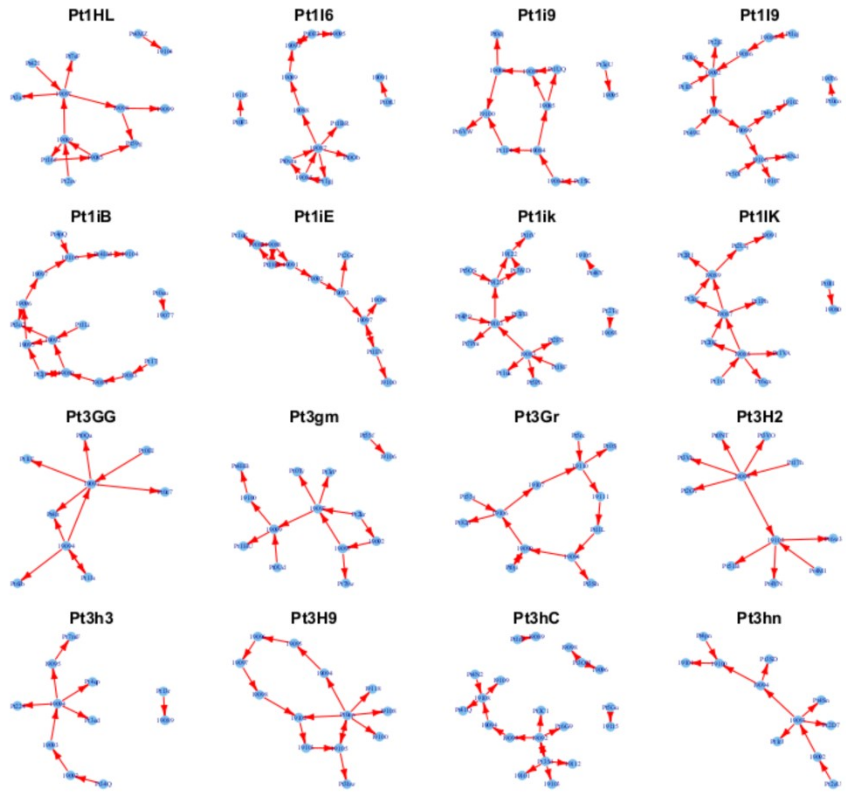


**Figure s01** The locations with 7 times as source of transmission (examples)

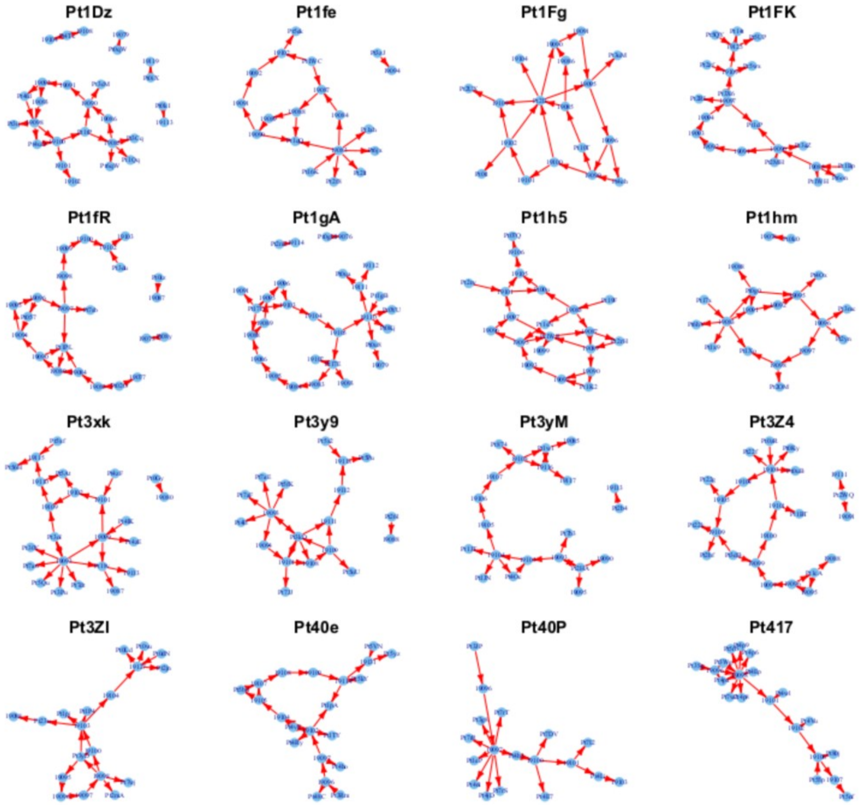


**Figure s02** The locations with 14 times as source of transmission (examples)

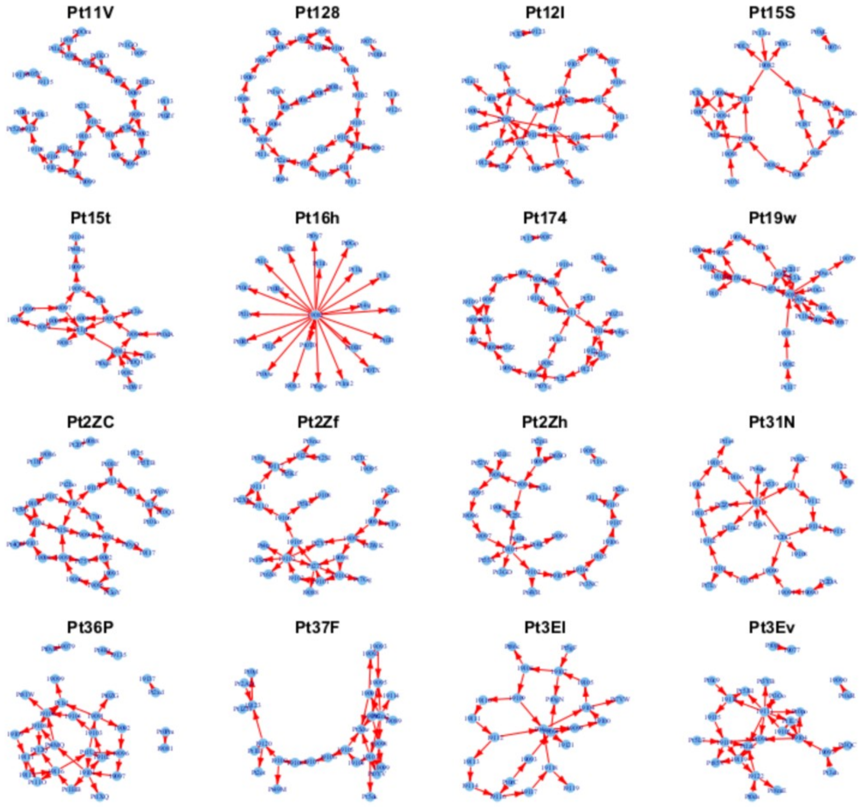


**Figure s03** The locations with 21 times as source of transmission (examples)

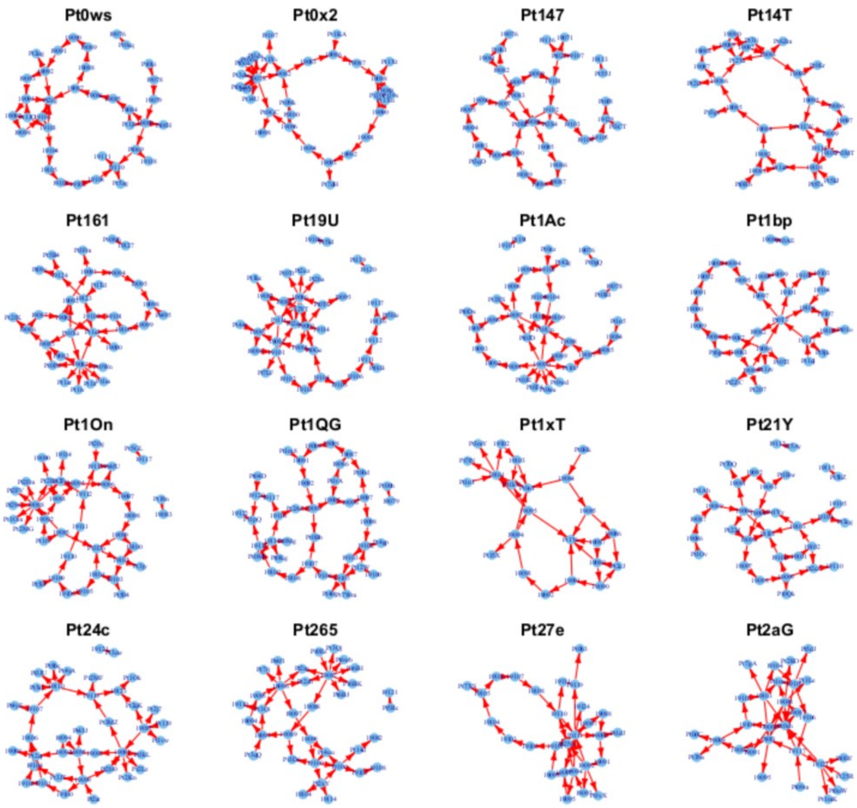


**Figure s04** The locations with 28 times as source of transmission (examples)

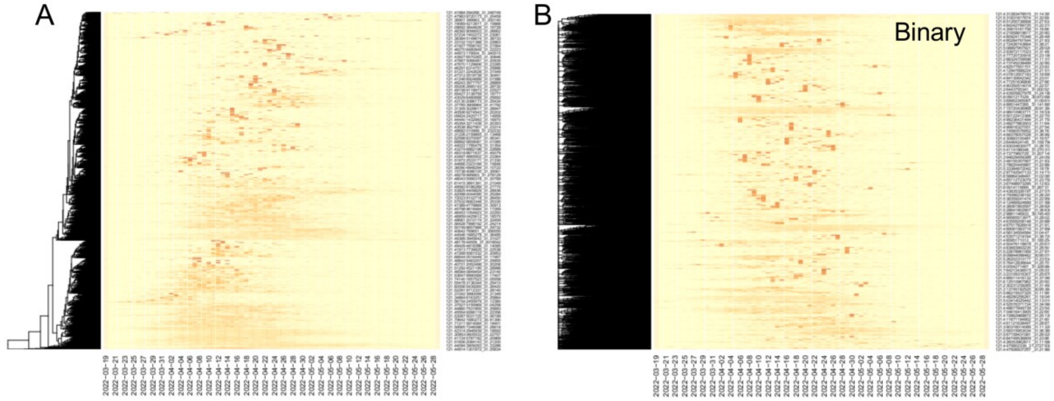


**Figure s05** The results of cluster of the repeatedly reported sample locations in chronologic sequence.

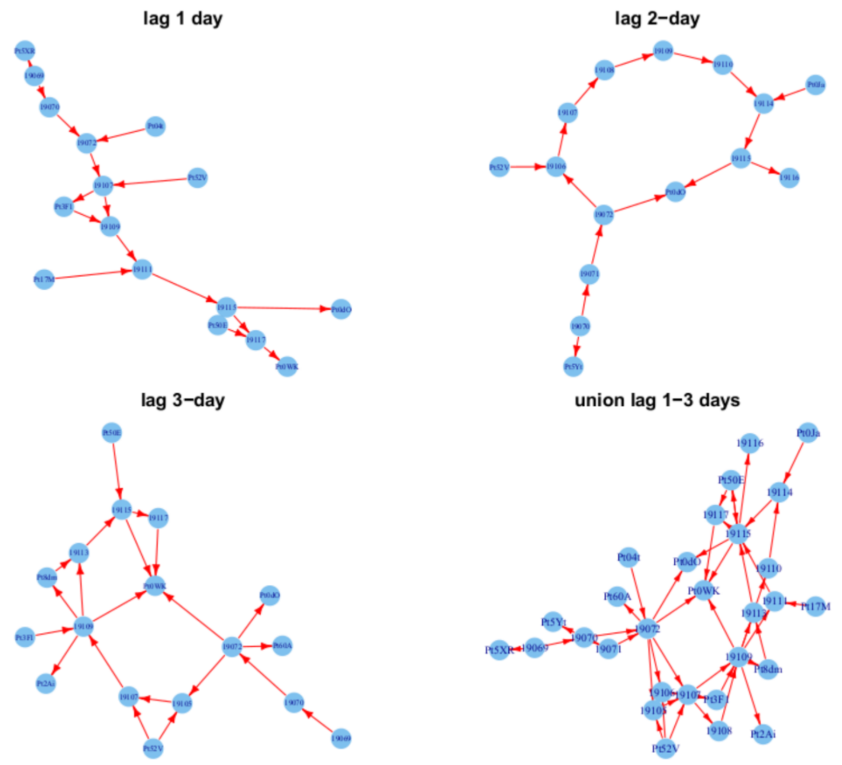


**Figure s06** Example of united point-wise transmission pattern of one to three days lag based upon nearest neighbour assumption

**Supplement B: Session information in R environment**

R version 4.0.1 (2020-06-06)

Platform: x86_64-w64-mingw32/x64 (64-bit)

Running under: Windows 7 x64 (build 7601) Service Pack 1

attached base packages:

[1] stats graphics grDevices utils datasets methods base

other attached packages:

[1] broom_0.7.9 rgdal_1.5-31 plyr_1.8.6 dplyr_1.0.0

[5] zoo_1.8-8 rgeos_0.5-9 maps_3.4.0 cluster_2.1.0

[9] ggplot2_3.3.2 igraph_1.2.5 maptools_1.0-2 sp_1.4-2

[13] spatstat_2.3-4 spatstat.linnet_2.3-2 spatstat.core_2.4-2 rpart_4.1-15

[17] nlme_3.1-148 spatstat.random_2.2-0 spatstat.geom_2.4-0 spatstat.data_2.2-0

[21] rgl_0.108.3

loaded via a namespace (and not attached):

[1] tidyselect_1.1.0 xfun_0.30 purrr_0.3.4 splines_4.0.1

[5] lattice_0.20-41 colorspace_1.4-1 spatstat.utils_2.3-0 vctrs_0.3.1

[9] generics_0.0.2 htmltools_0.5.2 mgcv_1.8-31 rlang_0.4.12

```
[13] pillar_1.4.4          foreign_0.8-80       glue_1.4.1           withr_2.2.0
[17] lifecycle_0.2.0       munsell_0.5.0        gtable_0.3.0         htmlwidgets_1.5.1
[21] knitr_1.39            fastmap_1.1.0        Rcpp_1.0.7           tensor_1.5
[25] backports_1.1.7       scales_1.1.1         jsonlite_1.7.0       abind_1.4-5
[29] deldir_1.0-6          digest_0.6.25        spatstat.sparse_2.1-1 polyclip_1.10-0
[33] grid_4.0.1            tools_4.0.1          magrittr_1.5         goftest_1.2-3
[37] tibble_3.0.1          tidyr_1.1.0          crayon_1.3.4         pkgconfig_2.0.3
[41] ellipsis_0.3.1        Matrix_1.2-18        R6_2.4.1             compiler_4.0.1
```

**Supplement C: Source code of user-defined function**

#The following code to generate the point-based chronological graph by using library(igraph) and library(zoo);

#To generate the graph by user-defined rules to get the dataframe containing Point Name, Date, From and To as indicated in toy dateset(Figure 5A);

```
chronBPts<-function(dfnm, varnm, datev, ppt, lag=1, realR=TRUE, plt=TRUE, ...){
    vln<-length(varnm)
    if (vln==1){            #for only From or To separately
  subdf01<-subset(x=dfnm, dfnm[,varnm[1]] %in% ppt)          #From points available
  subdf01<-subdf01[!duplicated(subdf01),]              #remove redundant records
        if (varnm=="From"){
     selfpi<-which(subdf01[, "From"]==subdf01[,"To"]) #date of self-propagation
     selfpdt<-as.integer(subdf01[selfpi, datev])+lag           #date propagation with lag
     }
  if (varnm=="To"){          #date of To plus lag
     subdf01[,datev]<-as.integer(subdf01[,datev])+lag
        selfpi<-which(subdf01[, "From"]==subdf01[,"To"])    #date of self-propagation
     selfpdt<-as.integer(subdf01[selfpi, datev])-lag           #date propagation with lag
     }
    ptsrc<-sort(as.integer(base::union(subdf01[,datev],selfpdt)))
        if (length(ptsrc)>1){
     ptsdt<-data.frame(zoo::rollapply(ptsrc, 2, function(x) {x}))
            names(ptsdt)<-c("From", "To")
            }
        else {
          ptsdt<-data.frame(From=ptsrc, To=ptsrc+lag)
            }
```

```
ptsdt<-ptsdt[ptsdt$From!=ptsdt$To,]                    #remove self connection
        grfdf<-data.frame(From=subdf01[,"From"], To=subdf01[,"To"])                    #same dim grfdf and
subdf01
    grfdf$From[which(grfdf$From==ppt)]<-subdf01[which(grfdf$From==ppt),datev]    #replace ppt with date

                if (varnm=="To"){
            grfdf$To[which(grfdf$To==ppt)]<-subdf01[which(grfdf$To==ppt),datev]    #replace ppt with date
            }
        for (ti in 1:dim(grfdf)[1]){                   #replace the To ppt with next date self-propagation
            if (grfdf$To[ti]==ppt){
                    grfdf$To[ti]<-ptsrc[match(grfdf$From[ti], ptsrc)+1]            #the next in date sequence
                    }
                }
        ptsdf<-rbind(ptsdt, grfdf)
        ptsdf<-ptsdf[ptsdf$From!=ptsdf$To,]              #redundant day-to-day transmission
                ptsdf<-ptsdf[!duplicated(ptsdf),]
        ptsdf<-ptsdf[order(ptsdf$From),]
                }

    if (vln>1){                                 #for both From and To combined
        if (!all(varnm==c("From", "To"))) stop("To provide From, To!")
        subdf01<-subset(x=dfnm, dfnm[,varnm[1]] %in% ppt)                    #From points available
    subdf01<-subdf01[!duplicated(subdf01),]                         #remove redundant records
                subdf02<-subset(x=dfnm, dfnm[,varnm[2]] %in% ppt)              #To points available
                subdf02<-subdf02[!duplicated(subdf02),]                        #remove redundant records
        subdf02[, datev]<-subdf02[, datev]+lag                          #To subset date plus lag
                selfpi<-which(subdf01[,"From"]==subdf01[,"To"])          #date of self-propagation
                selfpdt<-as.integer(subdf01[selfpi,datev])+lag                     #date propagation with lag
                ptsrc1<-sort(as.integer(base::union(subdf01[,datev], selfpdt)))                #From
                ptsrc2<-sort(as.integer(base::union(subdf02[,datev], selfpdt)))                #To
                ptsrc12<-sort(base::union(ptsrc1, ptsrc2))
                if (length(ptsrc1)>1 | length(ptsrc12)>1){
                    ptsdt1<-data.frame(zoo::rollapply(ptsrc1, 2, function(x) {x}))             #From To dates
            ptsdt12<-data.frame(zoo::rollapply(ptsrc12, 2, function(x) {x}))               #From To dates
                    names(ptsdt1)<-c("From", "To")                    #From
                    names(ptsdt12)<-c("From", "To")                   #From
```

```
            }
        else {
            ptsdt1<-data.frame(From=ptsrc1, To=ptsrc1+lag)
                ptsdt12<-data.frame(From=ptsrc12, To=ptsrc12+lag)
                }
        ptsdt1<-ptsdt1[ptsdt1$From!=ptsdt1$To,]                 #remove From self connection
    ptsdt1<-ptsdt1[!duplicated(ptsdt1),]
        ptsdt12<-ptsdt12[ptsdt12$From!=ptsdt12$To,]             #remove From self connection
        ptsdt12<-ptsdt12[!duplicated(ptsdt12),]

        grfdf1<-data.frame(From=subdf01[,"From"], To=subdf01[,"To"])            #same dim grfdf and
subdf01
    grfdf1$From[which(grfdf1$From==ppt)]<-subdf01[which(grfdf1$From==ppt),datev]          #replace From
ppt with date
        grfdf2<-data.frame(From=subdf02[,"From"], To=subdf02[,"To"])            #same dim grfdf and
subdf02
        grfdf2$To[which(grfdf2$To==ppt)]<-subdf02[which(grfdf2$To==ppt),datev]      #replace To
ppt with date
        print("Combined From and To subsets!"); print(rbind(grfdf1, grfdf2))

        for (ti in 1:dim(grfdf1)[1]){        #replace the To ppt with next date self-propagation
      if (grfdf1$From[ti]==ppt){
            grfdf1$From[ti]<-ptsrc1[match(as.integer(grfdf1$To[ti]), ptsrc1)]     #the next in date
sequence
            }
          }

        for (ti in 1:dim(grfdf2)[1]){        #replace the To ppt with next date self-propagation
      if (grfdf2$To[ti]==ppt){
            grfdf2$To[ti]<-ptsrc2[match(as.integer(grfdf2$From[ti]), ptsrc2)+1]             #the next in
date sequence
            }
          }
        if (realR) {        #use only the date of From
                ptsdf<-rbind(ptsdt1, grfdf1, grfdf2)        #reference dates two dfs
   }
```

```
        else {
          ptsdf<-rbind(ptsdt12, grfdf1, grfdf2)  #two dates two dfs
                }
      ptsdf$From[which(ptsdf$From==ppt)]<-ptsdf$To[which(ptsdf$From==ppt)]
        ptsdf$To[which(ptsdf$To==ppt)]<-ptsdf$From[which(ptsdf$To==ppt)]
        ptsdf<-ptsdf[ptsdf$From!=ptsdf$To,]           #redundant day-to-day transmission
        ptsdf<-ptsdf[!duplicated(ptsdf),] #unique data allow From equal to To means within lag repeat
        ptsdf<-ptsdf[order(ptsdf$From),]       #by order of From
        }

    #prepare to plot or not
    if (plt) {
        grfpt<-igraph::graph_from_data_frame(ptsdf)   #to generate DAG
      plot(grfpt, ...)
        return(grfpt)          #plot return graph
        }
    else {
      return(ptsdf)          #not plot return df
        }
}
#end

#Example
#for data from those in toy dataset dfName (name of dataframe);
#alternatively, varnm can be "From" or "To" or both(as in follows);
#data value, "Date1" the date of From;
#point name in ppt="Pt001";
#time lag in lag=1 or other integer;
chronBPts(dfnm=dfName, varnm=c("From","To"), datev="Date1", ppt="Pt001", lag=1)
```
